# Randomized Crossover Clinical Trial of Nicotinamide Riboside and Coenzyme Q10 on Metabolic Health and Mitochondrial Bioenergetics in CKD

**DOI:** 10.1101/2024.08.23.24312501

**Authors:** Armin Ahmadi, Ana P. Valencia, Gwénaëlle Begue, Jennifer E. Norman, Sili Fan, Blythe P. Durbin-Johnson, Bradley N. Jenner, Matthew D. Campbell, Gustavo Reyes, Pankaj Kapahi, Jonathan Himmelfarb, Ian H. de Boer, David J. Marcinek, Bryan R. Kestenbaum, Jorge L. Gamboa, Baback Roshanravan

**Affiliations:** Department of Medicine, Division of Nephrology, University of California, Davis, CA, USA; Department of Medicine, Division of Metabolism, Endocrinology and Nutrition, University of Washington, Seattle, WA, USA; Kinesiology Department, California State University, Sacramento, CA, USA; Department of Internal Medicine, Division of Cardiovascular Medicine, University of California, Davis, CA, USA; Department of Biostatistics, School of Medicine, University of California, Davis, CA, USA; Department of Radiology, University of Washington, Seattle, WA, USA; The Buck Institute for Research on Aging, Novato, CA 94945, USA; Leonard Davis School of Gerontology, University of Southern California, Los Angeles, CA, USA; Department of Medicine, Division of Nephrology, Kidney Research Institute, University of Washington, Seattle, WA, USA; School of Medicine, Vanderbilt University, Nashville, TN, USA

**Keywords:** Nephrology, clinical trial, metabolism

## Abstract

**Background:** Mitochondria-driven oxidative/redox stress and inflammation play a major role in chronic kidney disease (CKD) pathophysiology. Compounds targeting mitochondrial metabolism may improve mitochondrial function, inflammation, and redox stress; however, there is limited evidence of their efficacy in CKD.

**Methods:** We conducted a randomized, double-blind, placebo-controlled crossover trial comparing the effects of 1200 mg/day of coenzyme Q10 (CoQ10) or 1000 mg/day of nicotinamide riboside (NR) supplementation to placebo in 25 people with moderate-to-severe CKD (eGFR <60mL/min/1.73 m^2^). We assessed changes in the blood transcriptome using 3’-Tag-Seq gene expression profiling and changes in pre-specified secondary outcomes of inflammatory and oxidative stress biomarkers. For a subsample of participants (n=14), we assessed lymphocyte and monocyte bioenergetics using an extracellular flux analyzer.

**Results:** The (mean±SD) age, eGFR, and BMI of the participants were 61±11 years, 37±9 mL/min/1.73m^2^, and 28±5 kg/m^2^ respectively. Of the participants, 16% had diabetes and 40% were female. Compared to placebo, NR-mediated transcriptomic changes were enriched in gene ontology (GO) terms associated with carbohydrate/lipid metabolism and immune signaling while, CoQ10 changes were enriched in immune/stress response and lipid metabolism GO terms. NR increased plasma IL-2 (estimated difference, 0.32, 95% CI of 0.14 to 0.49 pg/mL), and CoQ10 decreased both IL-13 (estimated difference, –0.12, 95% CI of –0.24 to –0.01 pg/mL) and CRP (estimated difference, –0.11, 95% CI of –0.22 to 0.00 mg/dL) compared to placebo. Both NR and CoQ10 reduced 5 series F2-Isoprostanes (estimated difference, –0.16 and –0.11 pg/mL, respectively; P<0.05 for both). NR, but not CoQ10, increased the bioenergetic health index (BHI) (estimated difference, 0.29, 95% CI of 0.06 to 0.53) and spare respiratory capacity (estimated difference, 3.52, 95% CI of 0.04 to 7 pmol/min/10,000 cells) in monocytes.

**Conclusion:** Six weeks of NR and CoQ10 improved in oxidative stress, inflammation, and cell bioenergetics in persons with moderate to severe CKD.

## Introduction

The pathophysiology of chronic kidney disease (CKD) includes an abnormal metabolism of lipids^1^, amino acids^2^, and carbohydrates^3^; chronic inflammation^4^, oxidative stress^5^, and insulin resistance^6^. There is a growing interest in the use of supplements, like nicotinamide riboside (NR) and coenzyme Q10 (CoQ10), that target mitochondrial metabolism for their potential effects on cardiometabolic health. However, limited published evidence addresses their effect on oxidative stress, inflammation, and mitochondrial metabolism in patients with CKD.

Nicotinamide adenine dinucleotide (NAD^+^) is a crucial cofactor and electron carrier involved in mitochondrial biogenesis, bioenergetics, and redox homeostasis^7^. Sufficient NAD^+^ levels are needed for various anabolic and catabolic pathways, including glycolysis, tricarboxylic acid (TCA) cycle, oxidative phosphorylation, beta-oxidation, and pentose phosphate pathway^8^. Prior studies have shown that CKD is associated with a reduction in de novo NAD^+^ biosynthesis contributing to metabolic perturbations (increased inflammation, mitochondrial dysfunction, and oxidative stress)^9^. Indeed, long-term (20 weeks) NR supplementation modulated inflammation and mitochondrial function and prevented diabetic kidney disease progression in mice^10^. Despite promising results from rodent studies, the impact of NAD^+^ supplements on systemic mitochondrial function in patients with CKD is lacking.

CoQ10 is a fat-soluble coenzyme that transfers electrons from complexes I and II to complex III within the inner mitochondrial membrane^11^. Depressed CoQ10 levels are associated with inefficient electron transport and increased ROS production^12^. Patients with advanced kidney failure treated with hemodialysis have lower plasma concentrations of CoQ10 biologically linked to increased oxidative stress and ameliorable with CoQ10 supplementation^13^.

We recently showed in a randomized crossover clinical trial of NR or CoQ10 (CoNR trial NCT03579693) that short-term (six-week) supplementation with NR or CoQ10 did not improve the primary outcomes of physical endurance (VO_2_ peak and total work efficiency) in patients with moderate to severe CKD. However, NR supplementation resulted in favorable changes in mitochondrial metabolism and plasma lipid profile, altering levels of TCA cycle intermediates metabolized by NAD^+^-dependent enzymes and leading to a broad systematic decrease in lipotoxic ceramides and glucosylceramides linked to inflammation and cardiometabolic risk^14^. We also demonstrated that CoQ10 supplementation in patients with CKD increased circulating free fatty acids and decreased complex triglycerides, suggesting improved beta-oxidation of fatty acids^14^.

The present study assesses the impact of NR or CoQ10 compared to placebo on pre-specified secondary endpoints of plasma markers of oxidative stress and inflammation in addition to lymphocyte and monocyte bioenergetics in patients with CKD enrolled in the CoNR trial. In an exploratory analysis, we further assess treatment-associated gene expression changes in the context of previously detected findings in the CoNR trial, providing deeper insight into mechanisms plausibly linked to metabolic disturbances in CKD.

## Methods

### Study population and design

Coenzyme Q10 and nicotinamide riboside in chronic kidney disease (CoNR) trial is a placebo-controlled, double-blind, randomized, cross-over trial with patients randomly assigned to 6 treatment sequences, each comprising 3 randomly ordered treatment periods. Each participant underwent three treatment periods of 6 weeks duration (Clinicaltrials.gov ID NCT03579693) (**Supplemental Figures 1 and 2**) separated by a 1-week washout. The trial was conducted from November 2018 to April of 2021. The active study treatments were 1000 mg/day of nicotinamide riboside (Niagen®) or 1200 mg/day of coenzyme Q10 (Tishcon Corp.). At all treatment periods, each participant took the same amounts of identical-looking tablets. Participants, study physicians, assessors, and study staff were blinded to the treatment and sequence allocation of participants. There was a minimum of 4 visits for data collection: one at the baseline and three post-treatment visits (one at the end of each treatment and washout period). A more detailed description of the study procedures, recruitment, population, and primary outcomes of the study has been previously reported^14^. The study was approved by the ethical review board of University of Washington (STUDY00004998).

### Pre-specified primary, secondary, and exploratory outcomes

The primary outcomes of the study were changes in physical exercise endurance measured by maximal aerobic capacity (VO_2_ peak) and total work efficiency measured using graded cycle ergometry testing. We have previously assessed the secondary outcome of changes in metabolic/lipid profile response using semi-targeted metabolomics and lipidomics profiling. In the present study, we assessed additional pre-specified secondary outcomes to test if NR and CoQ10 supplementation changes plasma inflammatory (IL-6 and CRP) and oxidative stress biomarkers (F2-isoprostanes). Additionally, we tested if NR or CoQ10 improved lymphocyte and monocyte bioenergetics (spare reserve capacity). As an exploratory endpoint, we assessed NR and CoQ10-induced gene expression changes by performing transcriptomics analysis of whole blood.

### RNA extraction and sequencing

RNA was extracted from blood collected in PAXgene tubes according to the manufacturer’s protocol, “Manual Purification of Total RNA from Human Whole Blood Collected into PAXgene Blood RNA Tubes” (PreAnalytiX, cat # 762164, pgs. 52-58).

We evaluated the changes in the gene expression profile from the extracted RNA using 3’-Tag-Seq Gene Expression Profiling as previously described^15^. Briefly, barcoded 3’Tag-Seq libraries were prepared using the QuantSeq FWD kit (Lexogen, Vienna, Austria) for multiplexed sequencing according to the manufacturer’s recommendations. The fragment size distribution of the libraries was verified via micro-capillary gel electrophoresis on a LabChip GX system (PerkinElmer, Waltham, MA). The libraries were quantified by fluorometry on a Qubit instrument (Life Technologies, Carlsbad, CA) and pooled in equimolar ratios. Sixty-two libraries were sequenced on one lane of an Aviti sequencer (Element Biosciences, San Diego, CA) with single-end 100 base pair reads. The sequencing generated more than 4 million reads per library. The library preparation and sequencing were carried out at the DNA Technologies and Expression Analysis Cores at the UC Davis Genome Center.

### Blood cytokines and F2-isoprostane measurements

Cytokines were measured using plasma samples. CRP was measured with a Beckman Coulter (USA) DxC chemistry analyzer. Plasma IL-10, IL-12, IL-13, IL-4, IL-2, IL-8 TNF-α, IL-6, and IFN-γ were measured using multiplex electroluminescence assays (Meso Scale Discovery, Rockville, MD, USA). For F2-isoprostane measurements, internal standard [^2^H_4_]-15-F_2T_-isoprostane was added to each plasma sample prior to C-18 and silica solid phase extraction, thin layer chromatography, and derivatization to penta-fluorobenzyl ester, trimethylsilyl ether derivative before measurement by LC-MS as described previously^16^.

### T-cell and monocyte isolation and extracellular flux analysis

Bioenergetics data was collected from a subset of CoNR trial participants (**Supplemental Figure 1**, **Supplemental Table 1**). Cell isolation and bioenergetics were conducted in blinded samples. The protocol for monocyte and lymphocyte separation has been reported previously^17^. A standard Cell Mito Stress using the Seahorse Biosciences XF24 extracellular flux analyzer was conducted per protocol^18^. Respiratory parameters were calculated for basal (OCR basal – OCR antimycin A), ATP-linked respiration (OCR basal – OCR oligomycin), proton leak (OCR oligomycin – OCR antimycin A), maximal uncoupled respiration (OCR FCCP – OCR antimycin A), spare respiratory capacity (OCR FCCP – OCR basal) and bioenergetic health index (BHI) (log [(ATP-linked respiration x spare respiratory capacity)/(proton leak x non-mitochondrial respiration]). Respiration rates were normalized to the number of cells plated per well.

### Statistical Analyses

Differential expression analyses were conducted using Dream^16^, which extends the limma-voom Bioconductor pipeline^19^ to accommodate linear mixed effects models. The model used in Dream included treatment, visit, treatment sequence, RNA extraction batch, and a random intercept for subjects. In this exploratory analysis unadjusted P-values are presented. Low expressed genes were filtered using the function filterByExpr in edgeR, leaving a total of 13,101 genes for analysis. Gene ontology (GO) enrichment analyses of DE results were conducted using Kolmogorov-Smirnov tests, as implemented in the Bioconductor package topGO. Analyses were conducted for the biological process (BP), molecular function (MF), and cellular component (CC) GO ontologies. In addition to GO analysis, we performed parallel gene pathway analyses using KEGG and Reactome.

Linear mixed-effect models using log-transformed inflammatory biomarkers were conducted to detect differences across the three groups: placebo, CoQ10, and NR. To quantify the magnitude of treatment effect on the plasma inflammatory biomarkers, we determined the effect size using standardized mean difference. An effect size of 0.5 is considered a medium effect and an effect size above 0.8 is considered a large effect. Similarly, treatment effects of NR and CoQ10 on plasma isoprostanes and immune cell bioenergetics were assessed using linear mixed effects modeling. To account for multiple comparison testing, we employed the Benjamini-Hochberg procedure. We also assessed the potential carryover and period effects using linear mixed-effect models and found negligible lingering supplementation effects from all treatment periods. A P-value<0.05 was considered significant for all analyses unless stated otherwise. Analyses were conducted in R version 4.2.2 ^20^.

## Results

*Characteristics of the study population.* A total of 25 participants with a mean age (± SD) of 61 ± 11.6 years and eGFR of 36.9 ± 9.2 mL/min per 1.73m^2^ completed the study (**Supplemental Figure 1**). The characteristics of the study participants at baseline are summarized in **Table 1**.

**Table 1.**
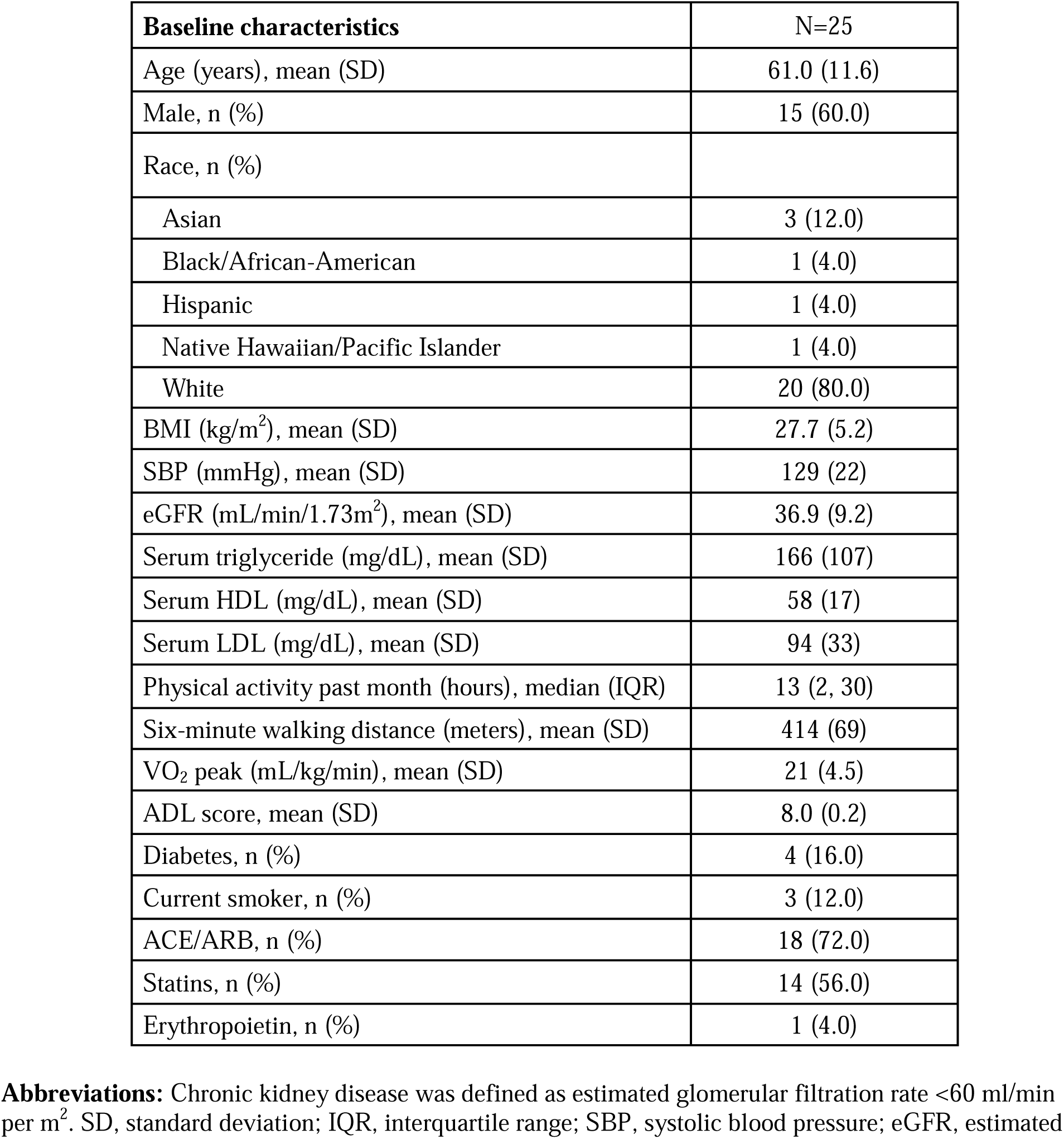
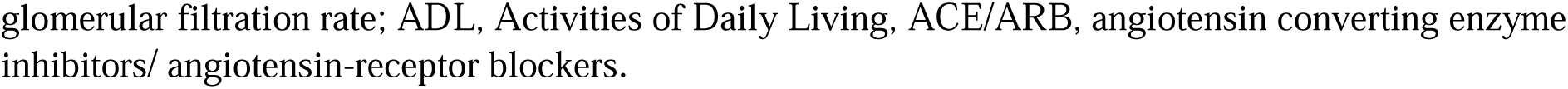
Participant characteristics of analytic population (n=25).

| **Baseline characteristics** | **N=25** |
| --- | --- |
| Age (years), mean (SD) | 61.0 (11.6) |
| Male, n (%) | 15 (60.0) |
| Race, n (%) |  |
| Asian | 3 (12.0) |
| Black/African-American | 1 (4.0) |
| Hispanic | 1 (4.0) |
| Native Hawaiian/Pacific Islander | 1 (4.0) |
| White | 20 (80.0) |
| BMI (kg/m<sup>2</sup>), mean (SD) | 27.7 (5.2) |
| SBP (mmHg), mean (SD) | 129 (22) |
| eGFR (mL/min/1.73m<sup>2</sup>), mean (SD) | 36.9 (9.2) |
| Serum triglyceride (mg/dL), mean (SD) | 166 (107) |
| Serum HDL (mg/dL), mean (SD) | 58 (17) |
| Serum LDL (mg/dL), mean (SD) | 94 (33) |
| Physical activity past month (hours), median (IQR) | 13 (2, 30) |
| Six-minute walking distance (meters), mean (SD) | 414 (69) |
| VO<sub>2</sub> peak (mL/kg/min), mean (SD) | 21 (4.5) |
| ADL score, mean (SD) | 8.0 (0.2) |
| Diabetes, n (%) | 4 (16.0) |
| Current smoker, n (%) | 3 (12.0) |
| ACE/ARB, n (%) | 18 (72.0) |
| Statins, n (%) | 14 (56.0) |
| Erythropoietin, n (%) | 1 (4.0) |

**Abbreviations:** Chronic kidney disease was defined as estimated glomerular filtration rate <60 ml/min per m<sup>2</sup>. SD, standard deviation; IQR, interquartile range; SBP, systolic blood pressure; eGFR, estimated
glomerular filtration rate; ADL, Activities of Daily Living, ACE/ARB, angiotensin converting enzyme inhibitors/ angiotensin-receptor blockers.

*NR supplementation altered gene sets associated with metabolism and immune/stress signaling.* Differential gene expression (DGE) analysis comparing NR supplementation to placebo revealed that 503 (4%) out of 13,101 detected genes were altered with 291 up-regulated and 212 downregulated genes (**Figure 1A**). The top three most up-regulated DGEs (by fold change) were AREG, RAP1GAP, and EGR3, with log2 fold changes of 1.5, 1.3, and 1.2, respectively. The top 3 most downregulated genes were MYBL2, MMP8, and CLIC1 with log-fold changes of –0.9, – 0.8, and –0.7, respectively. Using gene ontology (GO) analysis, a total of 185 biological processes and 55 molecular function terms were altered with NR supplementation. NR supplementation resulted in the alteration of genes with various GO terms (top 20 BP and MF) relating to carbohydrate metabolism (carbohydrate phosphorylation, glucose binding, and glucose transmembrane transporter activity), lipid metabolism (regulation of long-chain fatty acid import, LDL binding, and fatty acid binding), and immune and stress response (immunoglobulin production, LPS-mediated response, and regulation of IL-1 production). NR gene response was also enriched in GO terms related to mitochondrial function, including “regulation of mitochondrial membrane potential” and “proton-transporting ATPase activity” (**Figure 1B and Supplemental Figure 3A**).

**Figure 1.**
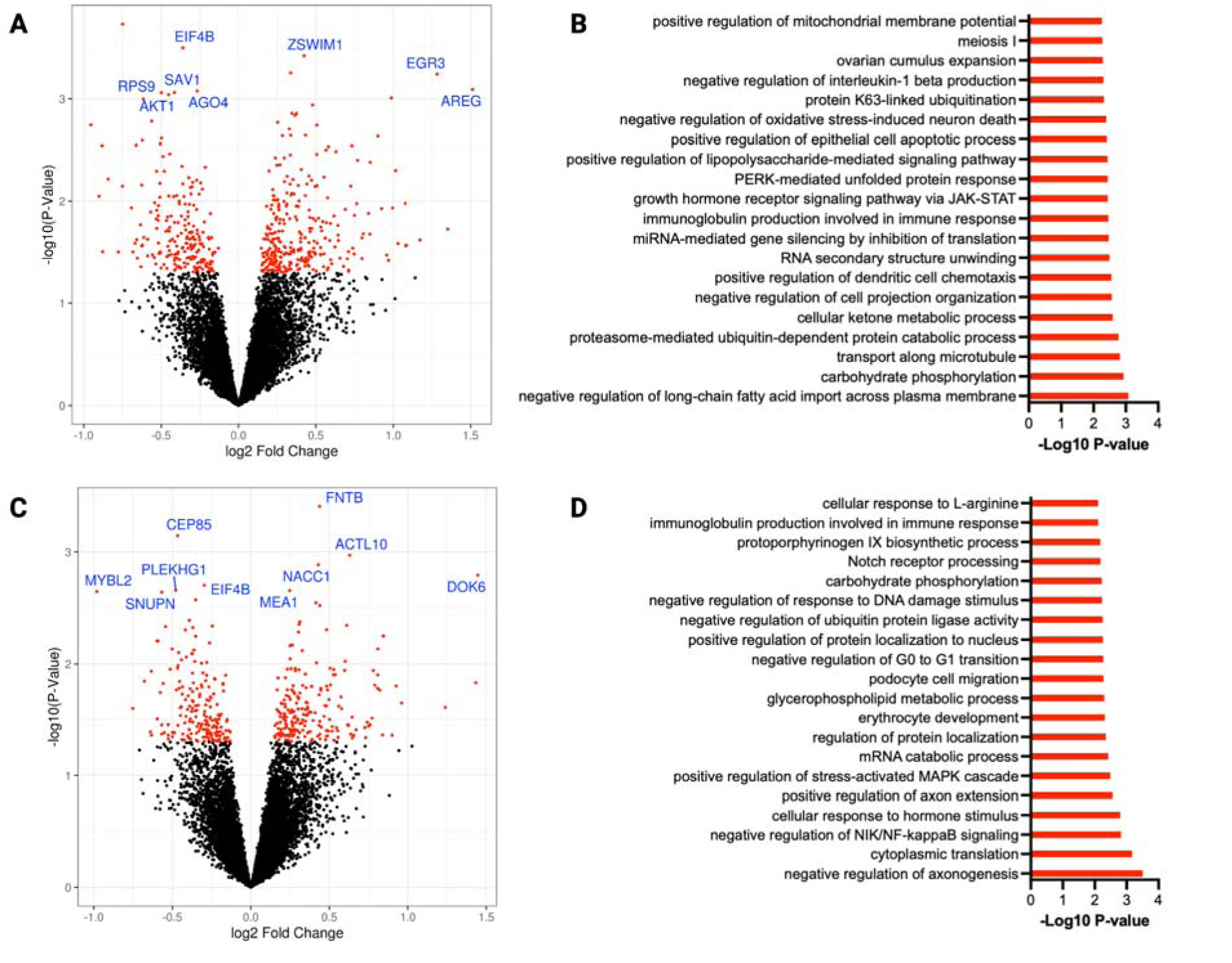
The impact of six weeks of NR and CoQ10 supplementation on whole blood transcriptomics profile in CKD. Volcano plot of differentially expressed genes comparing A) NR vs placebo and C) CoQ10 vs placebo illustrated by plotting the fold change of gene expression (Log2, x axis) against P-value for differential gene expression (–Log10, y axis). Red dots represent genes that are significantly altered (P-value <0.05) post NR or CoQ10 supplementation. The top 10 genes by p-value are indicated with their gene symbol. Gene ontology analysis representing altered biological processes upon B) NR and D) CoQ10 supplementation. Gene Ontology analysis was performed using GSEA. Bars represent the P-value (–Log10). Top 20 altered terms are shown. N=25

*CoQ10 supplementation altered gene sets associated with protein/RNA metabolism, immune/stress signaling, and lipid metabolism.* Compared to placebo, CoQ10 supplementation altered 389 (3% of detected) genes with 187 up-regulated and 203 downregulated genes (**Figure 1C**). The most up-regulated DEGs compared to placebo were DOK66, CSMD2, and RAP1GAP with a log2 fold changes of 1.4, 1.4, and 1.2 respectively. The most downregulated DEGs post CoQ10 were MYBL2, PDK4-AS1, and MMP8 with log fold changes of –0.9, –0.7, and –0.6 respectively (**Figure 1C**). GO analysis revealed that CoQ10 supplementation altered genes related to 208 biological process terms and 49 molecular function terms. The top 20 enriched GO terms were involved in various processes, including immune system response and inflammation (regulation of NIK/NF-κB signaling, regulation of stress-activated MAPK cascade, immunoglobulin production, and cytokine activity) and glycerophospholipid metabolism (**Figure 1C and Supplemental Figure 3B)**.

*NR and CoQ10 supplementation differentially influenced circulating plasma inflammatory biomarker levels in CKD.* Treatment with NR, resulted in a significant rise in IL-2 levels relative to placebo (estimated mean difference of 0.32 pg/mL, 95% CI of 0.14 to 0.49, P<0.01). To a lesser degree, NR modestly decreased IL-13 levels (P=0.05) compared to placebo (**Figure 2 and Supplemental Table 2**). CoQ10 supplementation resulted in a significant decrease in plasma IL-13 and CRP levels (estimated mean difference of –0.12 pg/mL, 95% CI of –0.28 to 0.00, P=0.02 and –0.27 mg/dL, 95% CI of –0.49 to –0.05, P=0.04 respectively) (**Figure 2 and Supplemental Table 2**). After adjustment for multiple comparison testing, CRP was only marginally lower compared to placebo (q=0.08).

**Figure 2.**
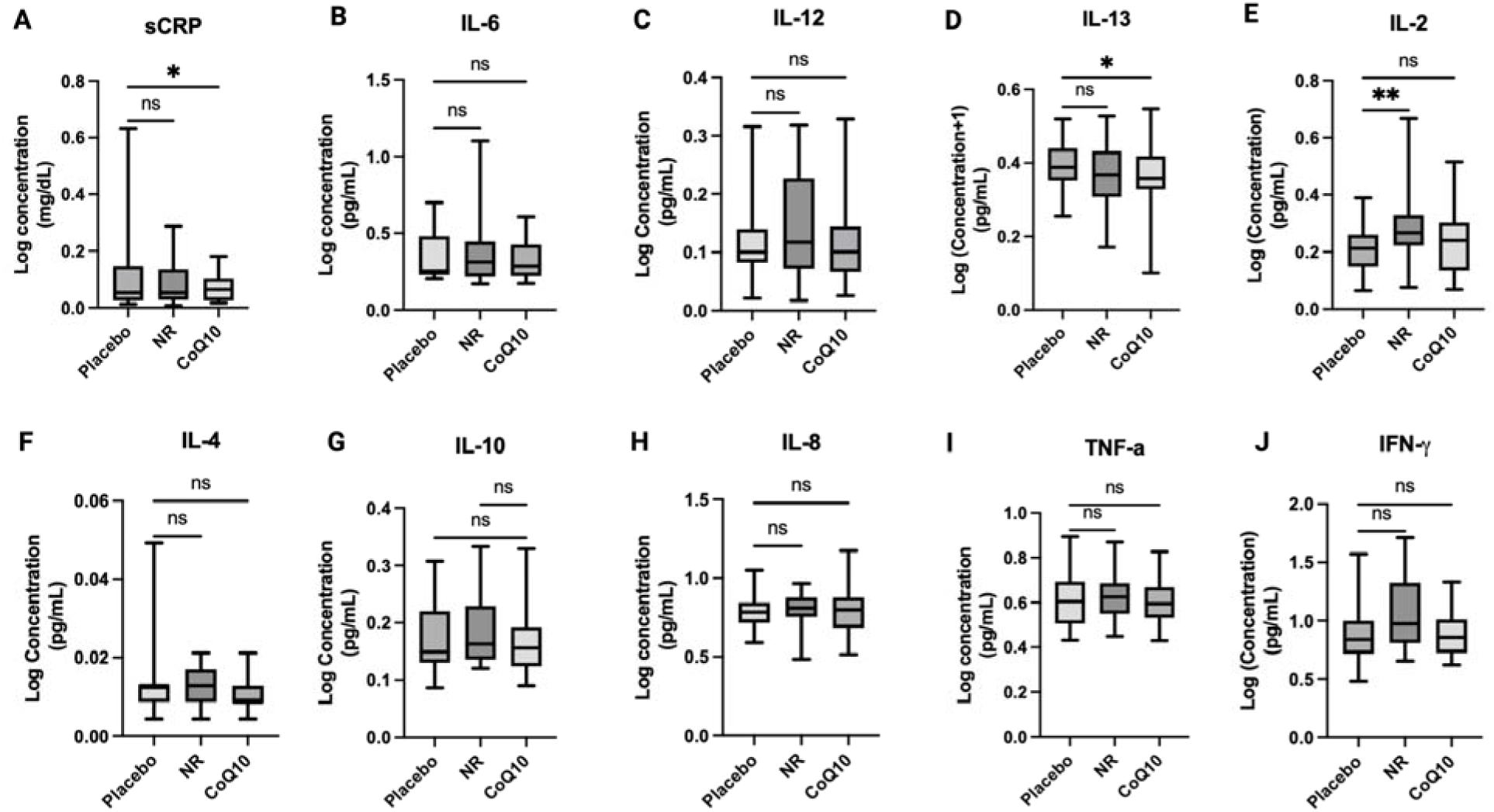
The impact of short-term NR and CoQ10 supplementation on circulating inflammatory biomarkers in CKD (n=25). A) sCRP, B) IL-6, C) IL-12, D) IL-13, E) IL-2, F) IL-4, G) IL-10, H) IL-8, I) TNF-α, J) IFNγ. The data is presented as log (concentration). The box plots represent median and IQR and the whiskers represent minimum and maximum values. Significance was determined using mixed effect modeling with a P < 0.05. Unadjusted P-values are shown. *P < 0.05, **P < 0.001.

*Both NR and CoQ10 supplementation resulted in a decrease in plasma markers of oxidative stress.* We assessed changes in three plasma isoprostanes as biomarkers of oxidative stress in response to NR and CoQ10 (**Figure 3**). NR significantly reduced total plasma F2-isoprostane levels compared to placebo. Both NR and CoQ10 supplementation resulted in reduction in 5 series isoprostanes but no changes in the 15 series isoprostanes compared to placebo (**Figure 3 and Supplemental Table 3**). After multiple comparison testing adjustment, the sum of 5 series (5-F2t plus 5-F2c) and 5-F2t isoprostanes alone were significantly lower post-NR and CoQ10 compared to placebo (q<0.05).

**Figure 3.**
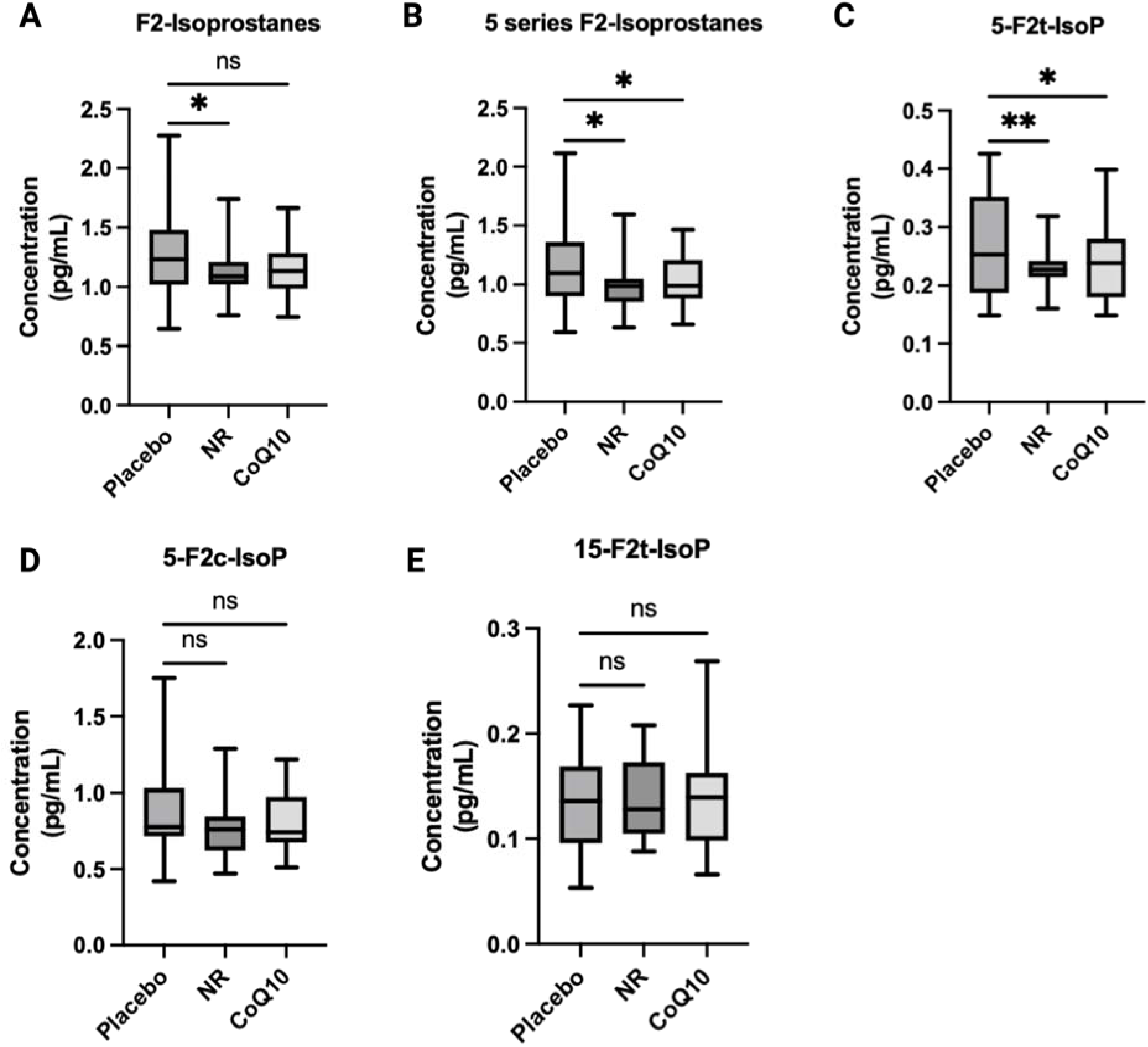
Changes in plasma markers of oxidative stress in response to NR and CoQ10 in CKD (n=25). A) F2 Isoprostanes, B) sum of 5 series F2-isoprostanes C) 5-F2t-Isoprostane, D) 5-F2c_Isoprostane, E) 15-F2t-Isoprostane. The box plots represent median and IQR and the whiskers represent minimum and maximum values. Unadjusted P-values are shown. Significance was determined using mixed effect modeling with a P < 0.05. *P < 0.05, **P < 0.001.

*NR supplementation, but not CoQ10 supplementation, increased respiratory capacity and BHI in monocytes*. Monocyte and T-cell bioenergetic profiles were available on 14 participants (**Supplemental Table 1**). NR led to significant changes in monocyte energetics compared to placebo (**Figure 4**, **Table 2, and Supplemental Figure 4**). NR supplementation significantly increased monocyte spare respiratory capacity by an estimated mean difference of 3.52 (95% CI of 0.04 to 7.0 pmol/min/10,000 cells, P=0.04) and BHI, an index representing mitochondrial function^21^ by an estimated mean difference of 0.29 (95% CI of 0.06 to 0.53, P=0.02) compared to placebo (**Figure 4 and Table 2**). Only BHI withstood multiple comparison testing adjustment (q<0.05). NR did not impact T-cell bioenergetics (**Supplemental Figure 5 and Table 2**). CoQ10 had no meaningful effect on monocyte or T-cell bioenergetics (**Table 2**).

**Figure 4.**
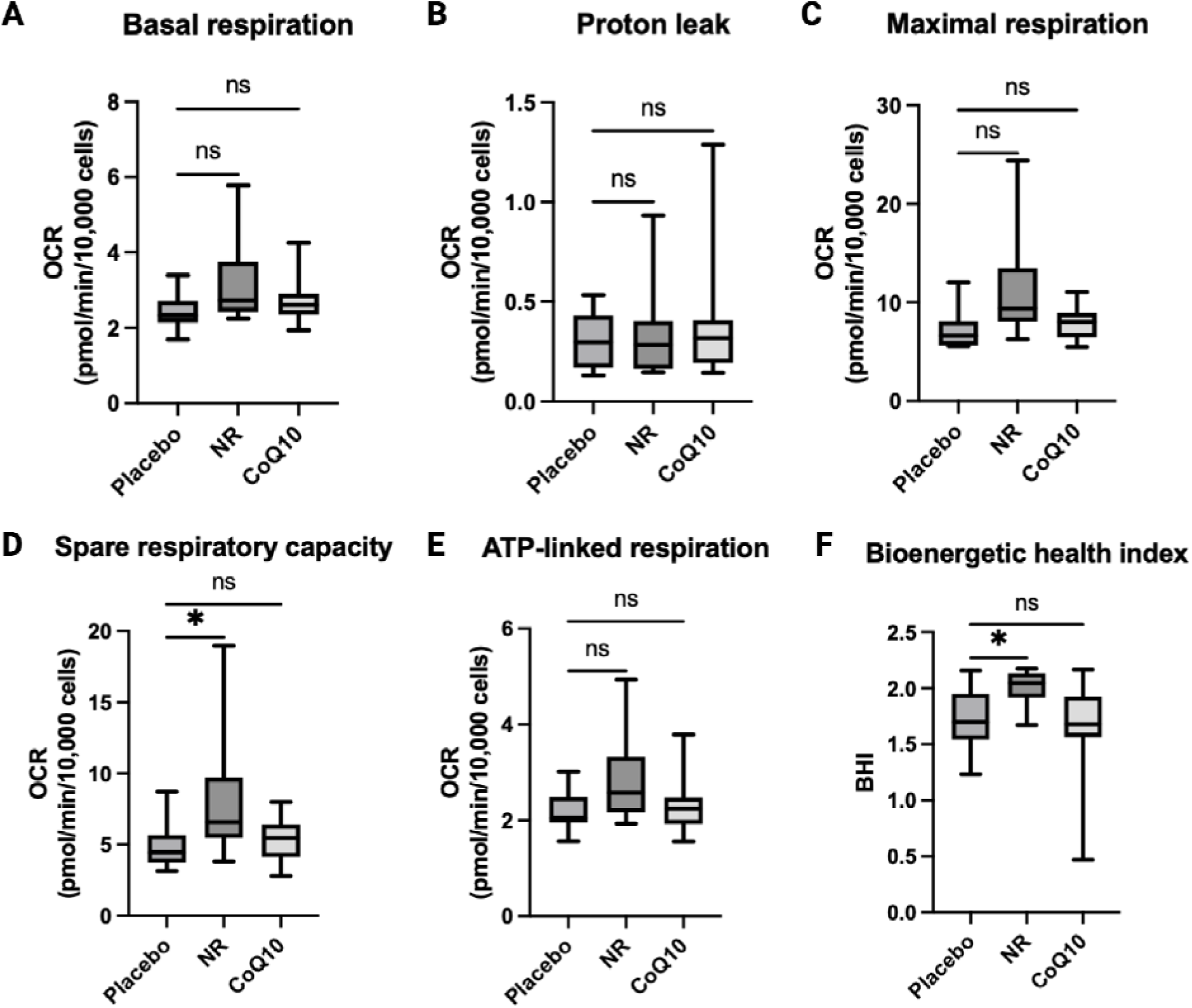
The effects of NR and CoQ10 on monocyte (CD14^+^) bioenergetics (n=14). Bioenergetic parameters include A) basal respiration, B) proton leak respiration, C) maximal respiratory capacity E) spare respiratory capacity, E) ATP-linked respiration, F) bioenergetic health index, calculated by the log [(ATP-linked respiration x spare respiratory capacity)/(proton leak x non-mitochondrial respiration]. The box plots represent median and IQR and the whiskers represent minimum and maximum values., *P<0.05 compared to placebo.

**Table 2.** Summary of secondary outcomes of monocyte and lymphocyte mitochondrial bioenergetics comparing NR and CoQ10 with placebo. Linear mixed effects modeling was used to estimate treatment effects compared to placebo. Unadjusted P-values, mean differences, and 95% CIs are shown. The comparisons that withstood multiple comparison adjustment are shown in bold.

| Endpoint | Mean Placebo (SD) | NR vs placebo (95% CI) | NR vs placebo P-value | CoQ10 vs placebo (95% CI) | CoQ10 vs placebo P-value |
| --- | --- | --- | --- | --- | --- |
| Secondary outcomes |  |  |  |  |  |
| Monocyte basal respiration, pmol/min/10,000 cells | 2.4 (0.5) | 0.79 (-0.14 to 1.7) | 0.08 | 0.24 (-0.11 to 0.60) | 0.15 |
| Monocyte proton leak, pmol/min/10,000 cells | 0.31 (0.14) | 0.02 (-0.12 to 0.16) | 0.73 | 0.05 (-0.18 to 0.29) | 0.60 |
| Monocyte maximal respiration, pmol/min/10,000 cells | 7.2 (1.9) | 4.32 (-0.01 to 8.64) | 0.05 | 0.74 (-0.61 to 2.10) | 0.24 |
| Monocyte spare reserve capacity, pmol/min/10,000 cells | 4.7 (1.5) | 3.52 (0.04 to 7.00) | 0.04 | 0.50 (-0.57 to 1.60) | 0.31 |
| Monocyte ATP-link respiration, pmol/min/10,000 cells | 2.2 (0.42) | 0.75 (-0.12 to 1.61) | 0.08 | 0.16 (-0.21 to 0.53) | 0.35 |
| Monocyte BHI | 1.7 (0.26) | 0.29 (0.06 to 0.53) | **0.02** | -0.04 (-0.36 to 0.28) | 0.78 |
| Lymphocyte basal respiration, pmol/min/10,000 cells | 0.26 (0.1) | -0.03 (-0.08 to 0.02) | 0.25 | 0.00 (-0.05 to 0.06) | 0.94 |
| Lymphocyte proton leak, pmol/min/10,000 cells | 1.7 (0.5) | -0.14 (-0.45 to 0.18) | 0.35 | -0.17 (-0.38 to 0.05) | 0.11 |
| Lymphocyte maximal respiration, pmol/min/10,000 cells | 5 (2.5) | -0.27 (-1.64 to 1.10) | 0.65 | -0.71 (-1.70 to 0.28) | 0.14 |
| Lymphocyte spare reserve capacity, pmol/min/10,000 cells | 3.2 (2.1) | -0.14 (-1.25 to 0.97) | 0.78 | -0.54 (-1.40 to 0.29) | 0.17 |
| Lymphocyte ATP-link respiration, pmol/min/10,000 cells | 1.5 (0.43) | -0.11 (-0.39 to 0.17) | 0.41 | -0.17 (-0.36 to 0.01) | 0.07 |
| Lymphocyte BHI | 1.7 (0.34) | 0.04 (-0.15 to 0.23) | 0.62 | 0.01 (-0.34 to 0.36) | 0.94 |

## Discussion

Using a placebo-controlled, double-blind randomized cross-over trial, we showed that six-week supplementation with NR or CoQ10 improved metabolic outcomes. Specifically, NR treatment altered the expression of genes associated with lipid and carbohydrate metabolism and immune response, coinciding with a decrease in markers of oxidative stress and an increase in the bioenergetic health index of circulating monocytes. CoQ10 supplementation also altered genes associated with immune and stress signaling and lipid metabolism, coinciding with reductions in the concentration of plasma markers of oxidative stress and inflammatory cytokines (CRP and IL-13). These observations build on our previous findings from the current clinical trial showing improved systemic mitochondrial metabolism and lipid profile in response to NR and CoQ10 supplementation^14^. Overall, these findings suggest that short-term CoQ10 and NR supplementation result in distinct improvements in metabolic health risk factors in moderate to severe CKD.

We found NR treatment targeted metabolic perturbations associated with CKD, leading to improvements in genes related to carbohydrate and lipid metabolism, expanding on our published findings of NR induced changes in TCA cycle plasma metabolites and lipid profile in persons with CKD^14^. Enhanced carbohydrate metabolism with NR is further evidenced by concomitant increase in submaximal respiratory exchange ratio during exercise testing^14^. We also detected GO terms related to lipid metabolism post-NR. This supports our previous findings of NR-induced improvements in the plasma lipid profile with a decrease in plasma glycerolipids, sphingolipids and glycerophospholipids in CKD^14^. Our findings are consistent with another study of 12 subjects showing skeletal muscle tissue transcriptional changes in energy-producing metabolic pathways,^22^ and a meta-analysis of clinical studies showing NAD^+^ precursor supplementation improves lipid metabolism^23^. Overall, our multi-omics data highlights the potential beneficial impact of short-term NR supplementation on carbohydrate and lipid metabolism mechanistically linked to improvements in mitochondrial function, perhaps via their known activation of sirtuins^24^.

CKD is accompanied by metabolic disturbances, including dyslipidemia, elevated inflammatory and oxidative stress markers associated with disease progression, and increased risk of mortality^25^. Studies of CoQ10 in CKD demonstrate improvement in markers of oxidative stress ^13,26^, but the impact on immune response and lipid metabolism is unknown. We showed six weeks of CoQ10 supplementation altered genes related to cell stress response, immune response, and lipid metabolism, supporting our prior findings from this same clinical trial that CoQ10 supplementation improves lipid profile in CKD by improving β-oxidation and decreasing plasma medium– and long-chain triglycerides^14^. Other studies have reported CoQ10 supplementation improves lipid profile, oxidative stress and antioxidant capacity in both CKD and non-CKD population^27,28^. Mechanistic in vitro studies suggest CoQ10 induces fatty acid oxidation and suppresses adipogenesis via PPAR-α-dependent activation of AMPK signaling ^29,30^. CoQ10 has also been shown to mitigate endothelial oxidative stress by modulating lectin-like oxidized low-density lipoprotein receptor (LOX-1)-mediated ROS generation via the AMPK/PKC/NADPH oxidase signaling pathway^30^. Together, our previous lipidomics and current transcriptomic analyses confirm improved lipid profile and stress/immune response with CoQ10 suggesting potential biological mechanisms, including MAPK signaling, NF-κB signaling, and altered glycerophospholipid metabolism for further clinical investigation.

Clinical studies of NR indicate heterogeneity in inflammatory marker response. In this study, NR increased levels of IL-2, a proinflammatory cytokine predominantly produced by activated CD4+ T-cells that can promote effector cells like CD8+ and also the function, maintenance, and survival of regulatory T cells (Tregs)^31^. Low-dose IL-2 can promote Treg expansion^31^, and NAD+ regulates Treg fate in vitro^32^. The significance of NR-induced elevations in IL-2 on lymphocyte immunometabolism and Tregs proliferation in CKD remains unknown. Our findings contrast with another trial of NR showing general reduction of inflammatory biomarkers in whole blood or immune cells^33,34^. Another 21-day randomized controlled cross-over trial of NR supplementation in older adults similar to our study showed suppressed levels of circulating inflammatory cytokines such as IL-2, and IL-6^22^. However, a separate clinical trial of six weeks of NR at 1000mg/daily in 13 middle-aged overweight and obese individuals did not impact blood inflammatory biomarkers^22^. Overall, our findings indicate a relatively modest impact of NR on inflammatory biomarkers in moderate to severe CKD.

In contrast to NR, six weeks of CoQ10 resulted in a significant reduction in plasma IL-13 and CRP levels compared to placebo. Our GO analysis revealed a GO term enriched in “negative regulation of NF-κB signaling” as a top 20 altered term post CoQ10 supplementation. Together, our findings highlight reduction of key markers of systemic inflammation in response to short-term CoQ10 supplementation’ possibly through mitigating ROS activation of NF-κB signaling. Biologic plausibility of these changes is supported by in vitro studies showing that CoQ10 modulates anti-inflammatory effects via reduction of nuclear factor-κB (NF-κB) dependent gene expression^35^. NF-κB activation by ROS upregulates proinflammatory cytokines expression^36^.

However, clinical studies have reported discrepant efficacy of CoQ10 supplementation on markers of inflammation. A meta-analysis of nine randomized controlled trials showed that CoQ10 supplementation significantly reduced circulating TNF-α levels without impacting CRP and IL-6^37^. Another meta-analysis reported that CoQ10 supplementation (60-300 mg/day) led to a significant reduction in IL-6 with marginal impacts on CRP levels in a cohort of subjects with cardiovascular disease risk factors^38^. These clinical studies are limited in the number of patients and a heterogeneous population with considerable differences in dosage, duration, and study design.

Compared to placebo, both NR and CoQ10 reduced plasma markers of oxidative stress. Systemic oxidative stress is well documented in CKD^5^ and is associated with worsened disease outcomes, increased risk of cardiovascular disease, and mortality^39,40^. F2-isoprostanes are products of non-enzymatic arachidonic acid peroxidation and a robust marker of in vivo oxidative stress^41^. We measured the two most common regioisomers (5– and 15-series F2 isoprostanes) that form as a result of arachidonic acid oxidation^42^. We found a significant decrease in 5-, but not 15-, series F2-isoprostanes with NR and CoQ10 supplementation separately compared to placebo. Human studies investigating the impact of NAD^+^ precursor supplementation on oxidative stress are sparse. A randomized cross-over study of 12 young and 12 older adults showed that an NR supplementation (500 mg) decreased F2-isoprostanes only among the older group ^43^. A previous randomized, placebo-controlled trial of 65 hemodialysis patients also showed a significant reduction in plasma F2-isoprostanes at 1200mg/daily of COQ10 after four months compared to baseline^26^. Overall, our findings confirm and build on the existing literature that both, NR and CoQ10 have anti-oxidative properties through improving metabolic function and anti-oxidative action.

Six weeks of NR supplementation improved monocyte bioenergetics but not T-cell bioenergetics compared to placebo. Immune cell mitochondrial bioenergetic profiling captures the effect of physiologic stressors such as inflammation and ROS, making it a complementary indicator of multisystem mitochondrial function^44^. Peripheral blood mononuclear cells (PBMC) bioenergetics often correlate with skeletal muscle and cardiac bioenergetics; both are strong predictors of exercise performance in patients with CKD and are associated with cardiometabolic risk in healthy population^45–47^. Our finding is consistent with studies in patients with heart failure showing that NR response correlated with increased PBMC bioenergetics and a decreased PBMC inflammatory cytokines expression^33^ ^34^. Together, our findings suggest that NR-induced improvement in monocyte bioenergetics coincide with a reduction in markers of oxidative stress. Further investigation is needed to determine if NR-induced changes in monocyte bioenergetics are mirrored in the skeletal muscle and if combination of NR and exercise may synergistically improved muscle metabolic health and cardiometabolic health parameters.

Our study had notable strengths and limitations. First, we employed an efficient randomized controlled crossover trial design to investigate two different therapies known to target mitochondrial function. Second, we performed our analyses in a clinical population with a comprehensive assessment of metabolomics and lipidomics, allowing a deeper probe into the mechanism of action. Our study was not without limitations. First, the power calculations to determine sample size were based on the primary outcome (aerobic capacity; VO_2_ peak), not the secondary outcomes. Second, the supplementation period was only 6 weeks, which limited our ability to evaluate longer-term treatment effects. Given the small sample size and the exploratory nature of our transcriptomics assessment, we did not adjust for multiple comparison testing in our DGE analysis.

In conclusion, six weeks of NR and CoQ10 supplementation have distinct beneficial impacts on whole blood transcriptome, inflammatory cytokines, and oxidative stress. Given that both NAD^+^ and CoQ10 bioavailability decreases with aging and CKD, studies combining NR/CoQ10 and exercise are warranted to assess the impact of combination therapy on mitochondrial function, cardiometabolic health biomarkers, and exercise adaptation in CKD.

## Disclosure

Authors have no conflict of interest to disclose.

## Data Availability

All data produced in the present study are available upon reasonable request to the authors

## Acknowledgments

We thank all the study participants for being part of this trial. We wish to express our gratitude to Dr. Philip A. Kramer for his efforts in helping to develop the cellular mitochondrial energetics protocols. We also thank ChromaDex as the manufacturer and provider of Niagen for this study.

## Author contributions

The conceptualization was contributed by AA, BR, and APV. The methodology was contributed by AA, SF, BDJ, and BR. The formal analysis was conducted by AA, SF, and BDJ. The investigation was performed by BR, BRK, JLG, and DJM. Resources were contributed by BRK, BR, JLG, and DJM. Data curation was performed by AA, APV, SF, BJ, and BDJ. The original draft was written by AA, APV, and BR. The review was written and edited by AA, BR, GB, APV, JLG, JEN, MDC, GR, BRK, PK, JH, IHDB, DJM, and BRK. Visualization was contributed by AA, APV, and BDJ. Supervision was carried out by BR, BRK, and DJM. Project administration was contributed by BRK, DJM, IHDB, JLG, JH, and BR. Funding acquisition was contributed by BR, BRK, JLG, and DJM.

## Data sharing statement

A complete deidentified patient metadata supporting the findings in this study has been made available on Figshare (DOI: 10.6084/m9.figshare.26511202). Additional information will be made available to share upon request.

**Supplemental Figure 1.**
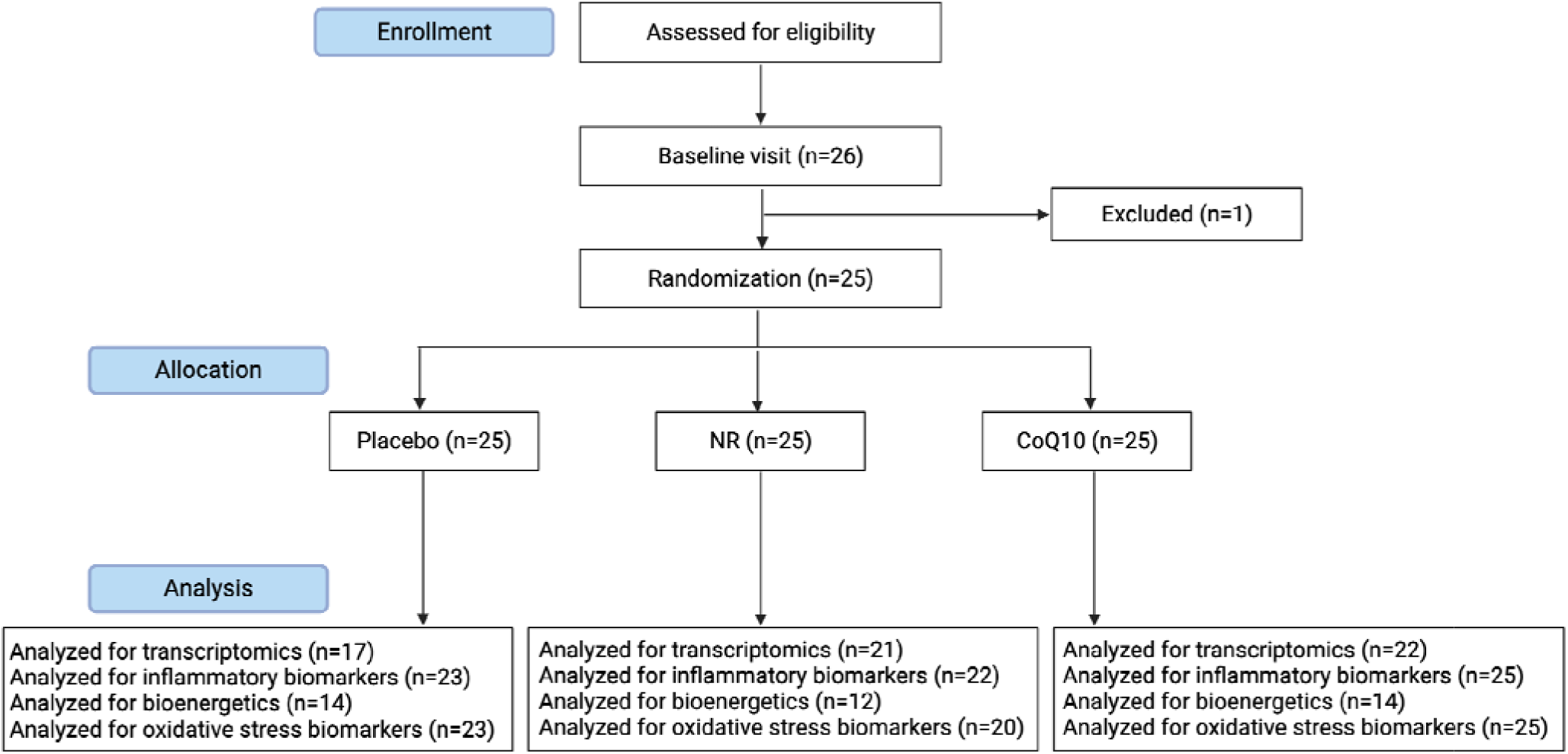
CONSORT diagram illustrating data completeness of the various outcomes in the Cross-over Randomized Controlled Trial of Coenzyme Q10 and Nicotinamide Riboside in CKD (CoNR Trial).

**Supplemental Figure 2.**
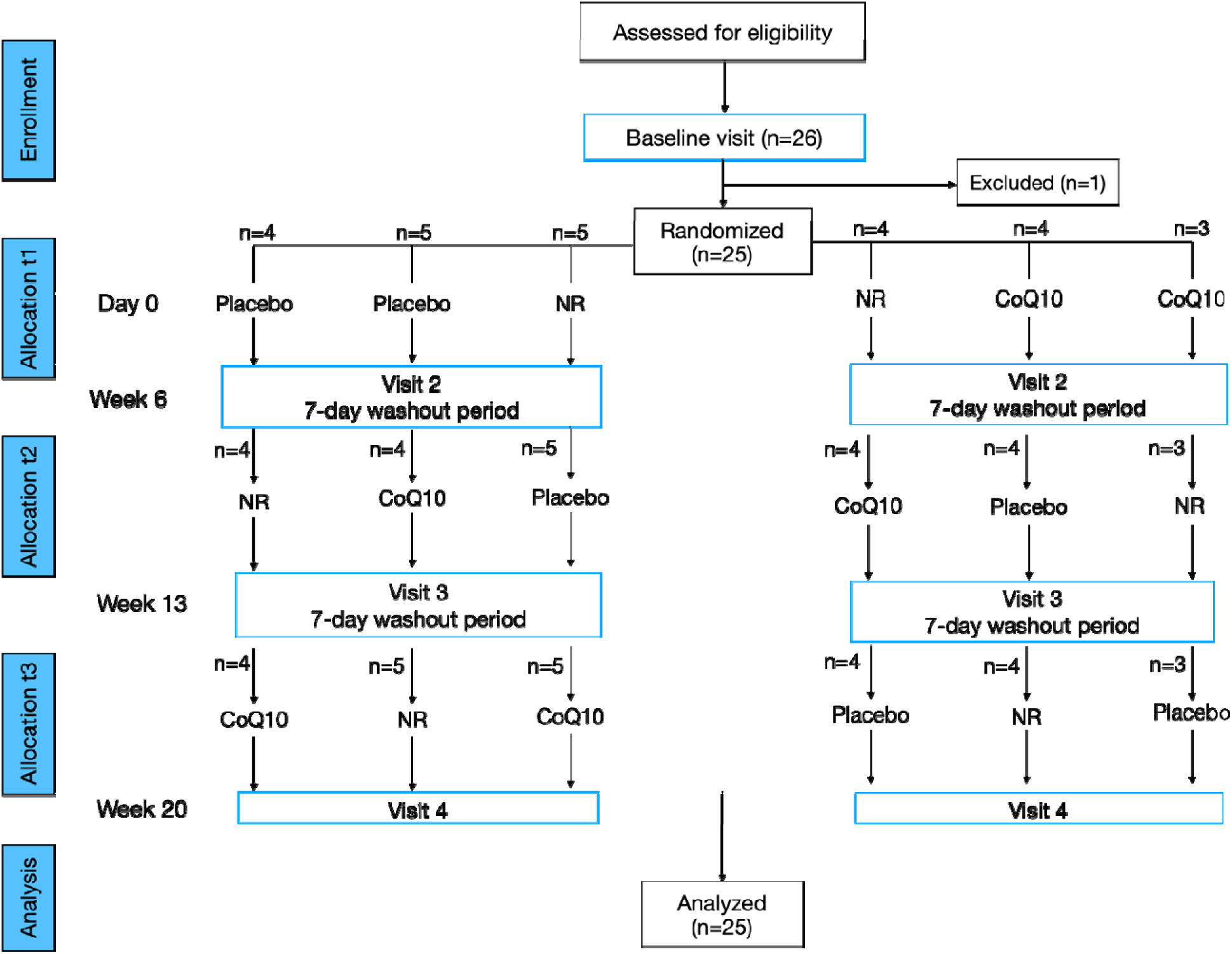
Study design and sample size of the Cross-over Randomized Controlled Trial of Coenzyme Q10 and Nicotinamide Riboside in CKD (CoNR Trial).

**Supplemental Figure 3.**
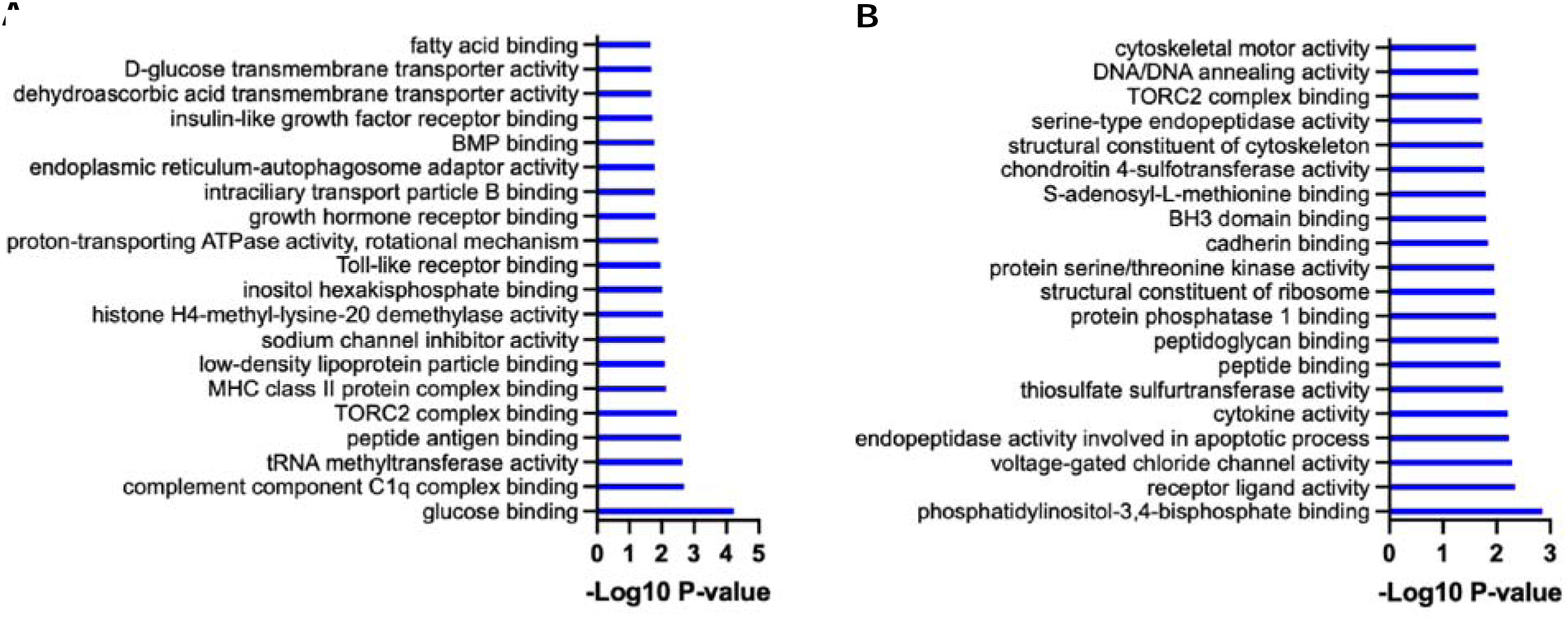
Gene ontology analysis representing altered molecular functions (mf) in whole blood (A) post NR and B) CoQ10 supplementation. Bars represent the P-value (–Log10). Top 20 terms are shown.

**Supplemental Table 1.**
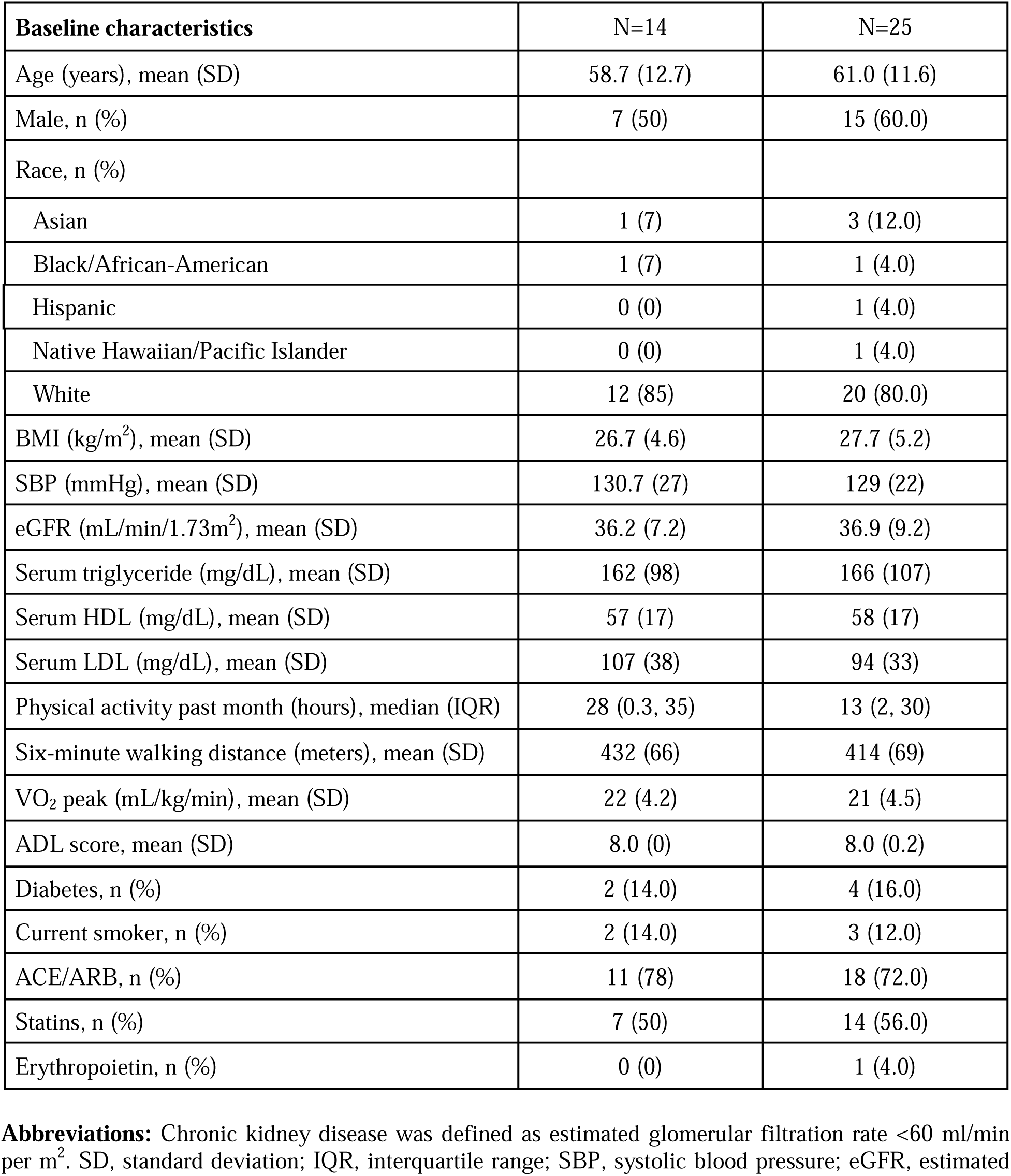
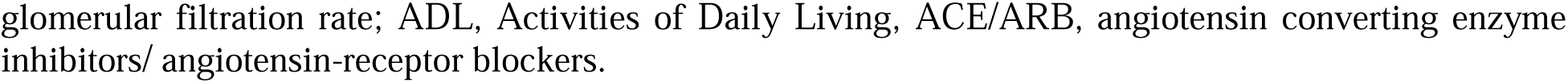
Sub-population characteristics of participants with monocyte and lymphocyte bioenergetics (n=14) compared to the entire cohort (n=25).

| Baseline characteristics | N=14 | N=25 |
| --- | --- | --- |
| Age (years), mean (SD) | 58.7 (12.7) | 61.0 (11.6) |
| Male, n (%) | 7 (50) | 15 (60.0) |
| Race, n (%) |  |  |
| Asian | 1 (7) | 3 (12.0) |
| Black/African-American | 1 (7) | 1 (4.0) |
| Hispanic | 0 (0) | 1 (4.0) |
| Native Hawaiian/Pacific Islander | 0 (0) | 1 (4.0) |
| White | 12 (85) | 20 (80.0) |
| BMI (kg/m<sup>2</sup>), mean (SD) | 26.7 (4.6) | 27.7 (5.2) |
| SBP (mmHg), mean (SD) | 130.7 (27) | 129 (22) |
| eGFR (mL/min/1.73m<sup>2</sup>), mean (SD) | 36.2 (7.2) | 36.9 (9.2) |
| Serum triglyceride (mg/dL), mean (SD) | 162 (98) | 166 (107) |
| Serum HDL (mg/dL), mean (SD) | 57 (17) | 58 (17) |
| Serum LDL (mg/dL), mean (SD) | 107 (38) | 94 (33) |
| Physical activity past month (hours), median (IQR) | 28 (0.3, 35) | 13 (2, 30) |
| Six-minute walking distance (meters), mean (SD) | 432 (66) | 414 (69) |
| VO<sub>2</sub> peak (mL/kg/min), mean (SD) | 22 (4.2) | 21 (4.5) |
| ADL score, mean (SD) | 8.0 (0) | 8.0 (0.2) |
| Diabetes, n (%) | 2 (14.0) | 4 (16.0) |
| Current smoker, n (%) | 2 (14.0) | 3 (12.0) |
| ACE/ARB, n (%) | 11 (78) | 18 (72.0) |
| Statins, n (%) | 7 (50) | 14 (56.0) |
| Erythropoietin, n (%) | 0 (0) | 1 (4.0) |

**Abbreviations:** Chronic kidney disease was defined as estimated glomerular filtration rate <60 ml/min per m<sup>2</sup>. SD, standard deviation; IQR, interquartile range; SBP, systolic blood pressure; eGFR, estimated
glomerular filtration rate; ADL, Activities of Daily Living, ACE/ARB, angiotensin converting enzyme inhibitors/ angiotensin-receptor blockers.

**Supplemental Table 2.** Summary of secondary outcome of plasma inflammatory biomarkers comparing NR and CoQ10 with placebo. Linear mixed effects modeling was used to estimate treatment effects compared to placebo. Unadjusted P-values, **effect sizes**, and 95% CIs are shown. The comparisons that withstood multiple comparison adjustment are shown in bold.

| Endpoint | NR vs placebo<br>Effect size<br>(95% CI) | NR vs<br>placebo<br>P-value | CoQ10 vs<br>placebo<br>Effect size<br>(95% CI) | CoQ10 vs<br>placebo<br>P-value |
| --- | --- | --- | --- | --- |
| sCRP, mg/dL | -0.32 (-0.91 to 0.29) | 0.13 | -0.45 (-1.05 to -0.17) | 0.04 |
| IL-6, pg/mL | 0.14 (-0.44 to 0.73) | 0.48 | -0.11 (-0.67 to 0.46) | 0.52 |
| IL-10, pg/mL | 0.23 (-0.36 to 0.81) | 0.10 | -0.1 (-0.67 to 0.46) | 0.57 |
| IL-12, pg/mL | 0.2 (-0.39 to 0.78) | 0.07 | 0.03 (-0.53 to 0.6) | 0.61 |
| IL-13, pg/mL | -0.35 (-0.97 to 0.27) | 0.05 | -0.37 (-0.1 to -0.25) | **0.02** |
| IL-2, pg/mL | 0.65 (0.04 to 1.24) | **<0.01** | 0.25 (-0.33 to 0.81) | 0.17 |
| IL-4, pg/mL | -0.13 (-0.71 to 0.46) | 0.41 | -0.37 (-0.94 to 0.02) | 0.16 |
| IL-8, pg/mL | 0.06 (-0.51 to 0.63) | 0.72 | 0.01 (-0.56 to 0.57) | 0.97 |
| TNF-α, pg/mL | 0.26 (-0.32 to 0.85) | 0.13 | 0.01 (-0.56 to 0.57) | 0.95 |
| IFN-γ, pg/mL | 0.55 (-0.06 to 1.15) | 0.12 | 0.06 (-0.51 to 0.62) | 0.80 |

**Supplemental Table 3.** Summary of secondary outcome of plasma inflammatory biomarkers comparing NR and CoQ10 with placebo. Linear mixed effects modeling was used to estimate treatment effects compared to placebo. Unadjusted P-values, mean differences, and 95% CI are shown. The comparisons that withstood multiple comparison adjustment are shown in bold.

| Endpoint | Mean placebo (SD) | NR vs placebo (95% CI) | NR vs placebo P-value | CoQ10 vs placebo (95% CI) | CoQ10 vs placebo P-value |
| --- | --- | --- | --- | --- | --- |
| F2 Isoprostanes, pg/mL | 1.26 (0.41) | 0.28 (0.00 to 0.25) | 0.05 | -0.02 to 0.23 | 0.09 |
| 5 series F2 Isoprostanes, pg/mL | 1.1 (0.4) | -0.16 (-0.28 to -0.03) | **0.02** | -0.11 (-0.22 to 0.00) | **0.04** |
| 5-F2t-IsoP, pg/mL | 0.27 (0.12) | -0.04 (-0.07 to -0.01) | **<0.01** | -0.03 (-0.06 to 0.00) | **0.03** |
| 5-F2c-IsoP, pg/mL | 0.86 (0.31) | -0.09 (-0.18 to 0.02) | 0.06 | -0.07 (-0.16 to 0.03) | 0.17 |
| 15-F2t-IsoP, pg/mL | 0.15 (0.05) | 0.00 (-0.02 to 0.02) | 0.78 | 0.00 (-0.02 to 0.02) | 0.96 |

**Supplemental Figure 4.**
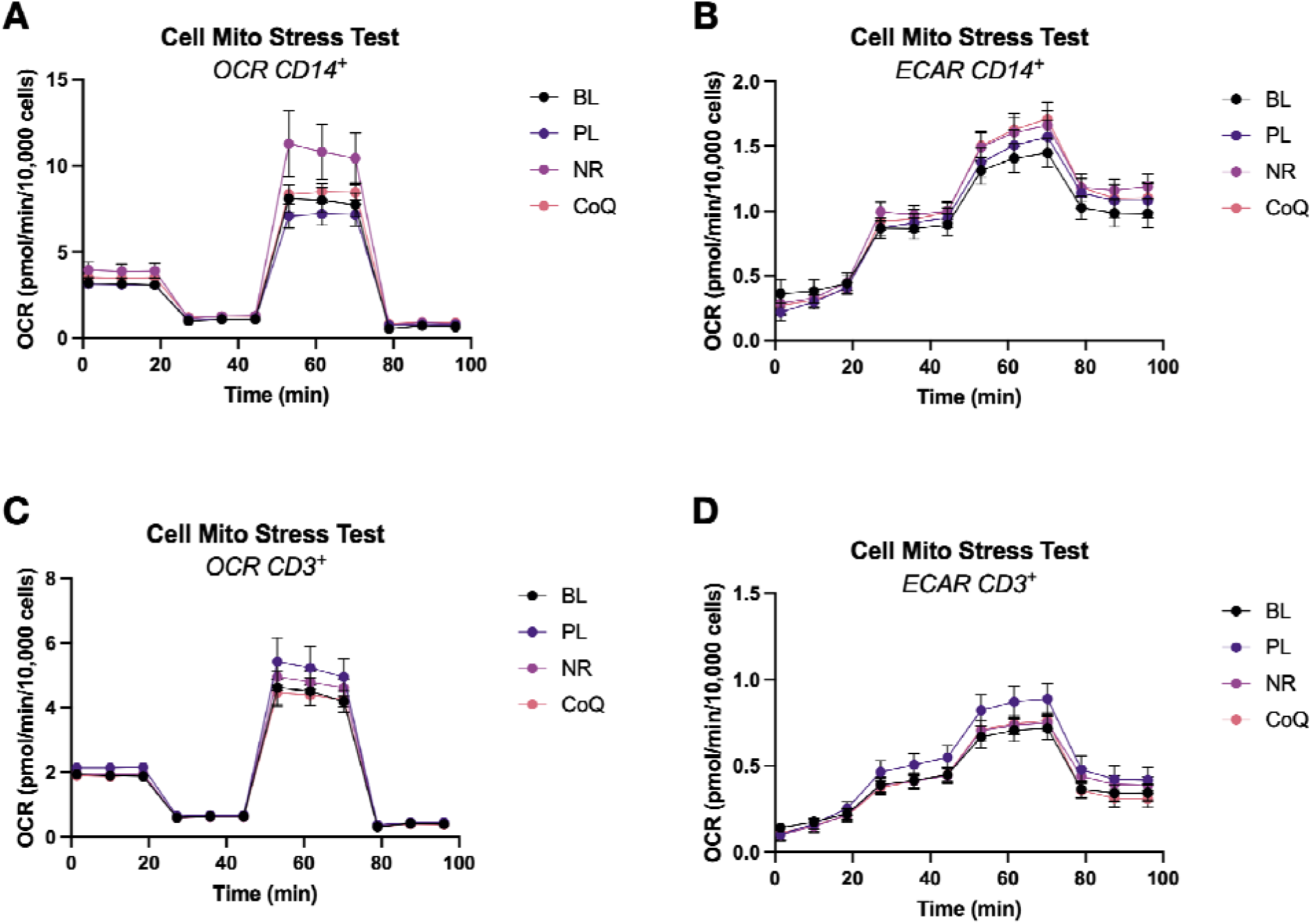
Oxygen consumption rates of A) monocytes and C) T-cells and extracellular acidification rates (ECAR) of B) monocytes and D) T-cells measured by an XFe24 Seahorse Analyzer (n=14). The *Cell Mito Stress* test involved an injection of 1μM oligomycin (OMY), followed by an injection of 0.6-1.0 μM carbonyl cyanide-4-(trifluoromethoxy)phenylhydrazone (FCCP), and an injection of 10μM antimycin A (AMA) in a XFe24 Analyzer. Data are normalized to cell number per well. Data expressed as mean±SEM.

**Supplemental Figure 5.**
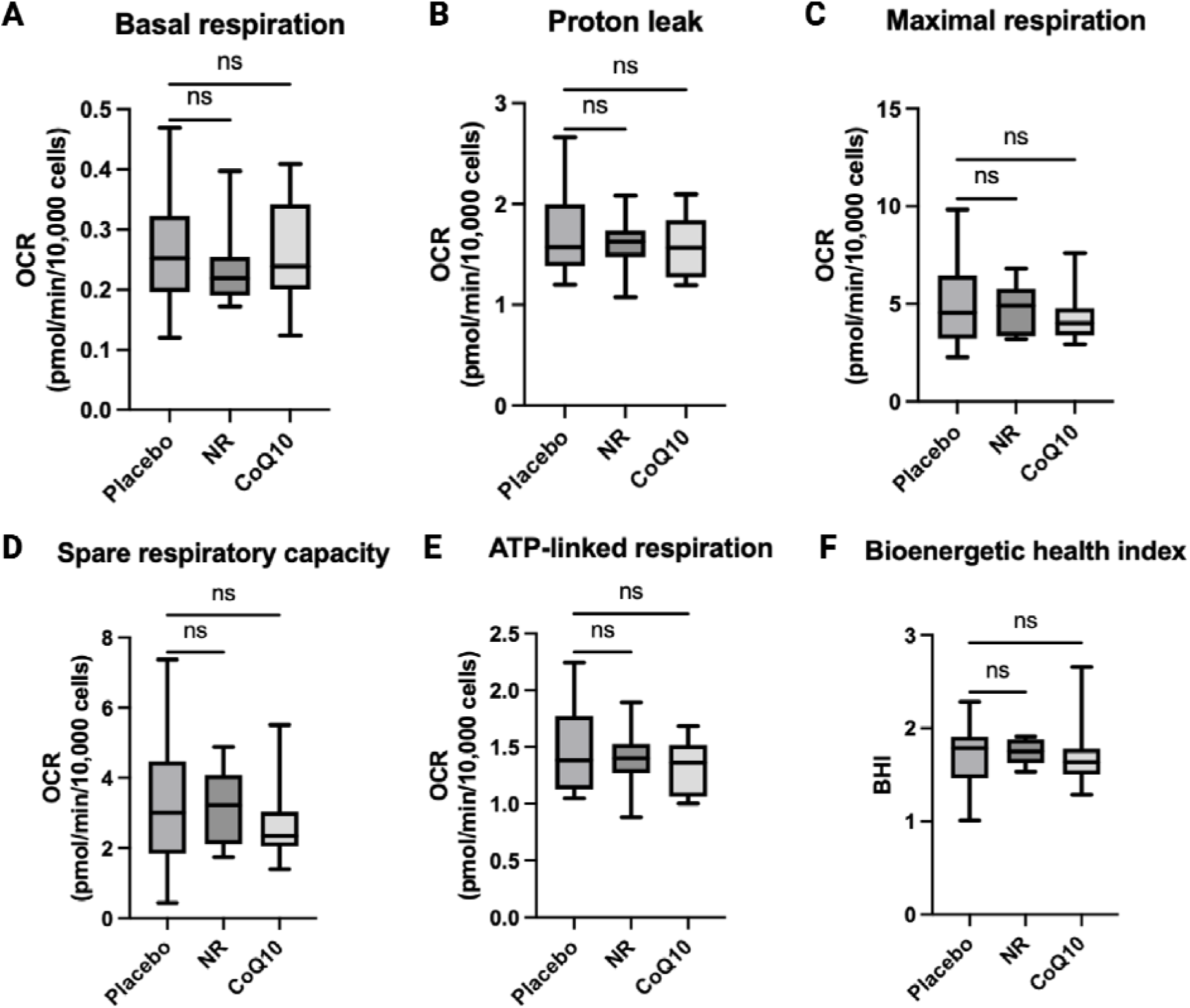
The effects of nicotinamide riboside (NR), and coenzyme Q 10 (CoQ10) on T-cell (CD3^+^) bioenergetics (n=14). Bioenergetic parameters include A) basal respiration, B) proton leak respiration, C) maximal respiratory capacity E) spare respiratory capacity, E) ATP-linked respiration, F) bioenergetic health index, calculated by the log [(ATP-linked respiration x spare respiratory capacity)/(proton leak x non-mitochondrial respiration]. The box plots represent median and IQR and the whiskers represent minimum and maximum values. *P<0.05 compared to placebo.

